# Enhancing SARS-CoV-2 Lineage surveillance through the integration of a simple and direct qPCR-based protocol adaptation with established machine learning algorithms

**DOI:** 10.1101/2024.08.09.24310239

**Authors:** Cleber Furtado Aksenen, Débora Maria Almeida Ferreira, Pedro Miguel Carneiro Jeronimo, Thais de Oliveira Costa, Ticiane Cavalcante de Souza, Bruna Maria Nepomuceno Sousa Lino, Allysson Allan de Farias, Fabio Miyajima

## Abstract

The emergence of the SARS-CoV-2 and continuous spread of its descendent lineages have posed unprecedented challenges to the global public healthcare system. Here we present an inclusive approach integrating genomic sequencing and qPCR-based protocols to increment monitoring of variant Omicron sublineages. Viral RNA samples were fast tracked for genomic surveillance following the detection of SARS-CoV-2 by diagnostic laboratories or public health network units in Ceara (Brazil) and analyzed using paired-end sequencing and integrative genomic analysis. Validation of a key structural variation was conducted with gel electrophoresis for the presence of a specific ORF7a deletion within the “BE.9” lineages. A simple intercalating dye-based qPCR assay protocol was tested and optimized through the repositioning primers from the ARTIC v.4.1 amplicon panel, which was able to distinguish between “BE.9” and “non-BE.9” lineages, particularly BQ.1. Three ML models were trained with the melting curve of the intercalating dye-based qPCR that enabled lineage assignment with elevated accuracy. Amongst them, the Support Vector Machine (SVM) model had the best performance and after fine-tuning showed ∼96.52% (333/345) accuracy in comparison to the test dataset. The integration of these methods may allow rapid assessment of emerging variants and increment molecular surveillance strategies, especially in resource-limited settings. Our approach not only provides a cost-effective alternative to complement traditional sequencing methods but also offers a scalable analytical solution for enhanced monitoring of SARS-CoV-2 variants for other laboratories through easy-to-train ML algorithms, thus contributing to global efforts in pandemic control.

## INTRODUCTION

The adaptability of viruses like SARS-CoV-2 through cumulative mutations denotes the dynamic interaction between pathogens and their environment. Mutations leading to structural modifications, such as insertions or deletions, are more likely to account for significant alterations in the biological behavior of the virus, ultimately fueling the emergence of variants with potential selective advantage and pathogenic profiles ^1–3^. This adaptive mechanism has been illustrated by the emergence of SARS-CoV-2 variants, which is known for its increased infectivity due to specific amino acid substitutions ^4,5^. The genetic diversity observed in RNA viruses, underscored by the continuous emergence of new mutations, highlights the evolving nature of these pathogens and the critical role of genomic surveillance in tracking these changes ^3,6,7^. The swift emergence and global proliferation of the Omicron variant (B.1.1.529) of SARS-CoV-2, along with its descendant subvariants, have heightened global apprehensions because of their extensive repertoire of distinctive genetic configurations and unprecedented transmission capabilities ^8,9^. Studies have illuminated the variant’s ability to outpace previous strains, such as the Delta variant, in terms of spread, leading to a considerable uptick in reinfection rates, affecting even those previously vaccinated or infected ^10–13^. The situation is compounded by the variant’s elusive severity profile compared with its predecessors, necessitating rigorous public health interventions ^14–16^. Given the dynamic nature of the virus, heightened emphasis on genomic surveillance is imperative to track and understand the emergence of new strains, enabling proactive measures to mitigate their spread and impact.

The global effort to monitor and control the spread of SARS-CoV-2 faces numerous challenges, including the economic and infrastructural disparities among countries. The challenges posed by NGS analyses, including high costs, lengthy response times, and its inaccessibility in economically disadvantaged regions, have spurred the scientific community to explore supplementary techniques ^17–19^. There has been a notable change toward integrating polymerase chain reaction (PCR) and computational algorithms into the genomic surveillance toolkit. These methods offer a more immediate and cost-effective capability for detecting specific genetic markers, thereby enhancing the efficiency and scope of pathogen surveillance efforts ^20,21^.

Among this landscape of innovation, the intercalating dye-based qPCR protocol has emerged as an important technique in the field of genetic surveillance. Distinguished by its capacity for real-time DNA amplification monitoring and low cost, this protocol has shown remarkable efficacy in pinpointing specific genetic markers. Its use not only marks a significant advanced strategy in the rapid identification of variants of concern (VOCs) but also in understanding the intricate dynamics of viral adaptations ^22,23^. The protocols’ insights into the ORF7a gene, particularly its role in immune modulation and interaction with host cells, underscore the complex interplay between viral genetics and host defenses, highlighting the importance of nuanced genetic surveillance in preparedness to the challenges of the COVID-19 pandemic and beyond ^3,24,25^.

The intercalating dye-based qPCR protocol is a low-cost assay technique, highly adaptable to near real time tool in the field of genomic surveillance, due to its steadfast deployment. This strategy can significantly improve both the speed and precision for target detections, proving reliable for the confirmation of key molecular signatures used for tracking the population dynamics and evolution of pathogens, such as SARS-CoV-2. Our work has highlighted the applicability of a lineage-defining genetic marker, a 244-base deletion within the ORF7a gene (27508 – 27751) characteristic marker of the Brazilian BE.9 lineage. We proposed this specific deletion could be informative and able to track in the spread of this lineage from September 2022 to May 2023 (https://gisaid.org/), underscoring the utility of a qPCR-based protocol in pinpointing the expansion of emerging variants and sublineages that pose new challenges to public health and vaccine efficacy. This seamless integration of computational analyses and a straightforward intercalating dye-based qPCR protocol represents a more direct and inclusive approach to monitoring viral evolution. It embodies the scientific community’s and public health policies in engaging in rapid response measures to monitor evolving pathogen variants, ensuring that public health strategies remain robust and responsive in the ongoing battle against immune escape and SARS-CoV-2 adaptability.

## RESULTS AND DISCUSSION

### Integrative Genomic Analysis and Categorization

SARS-CoV-2 genomic sequences with high-quality samples (horizontal coverage exceeding 90% and vertical coverage surpassing 100x) revealed a low depth region at position 27,508 – 27,751 of the ORF7a gene for previously classified as “BE.9” **(Figure 2A)**, when compared with classified as “non-BE.9”, particularly BQ.1 **(Figure 2B)**.

**Figure 1.**
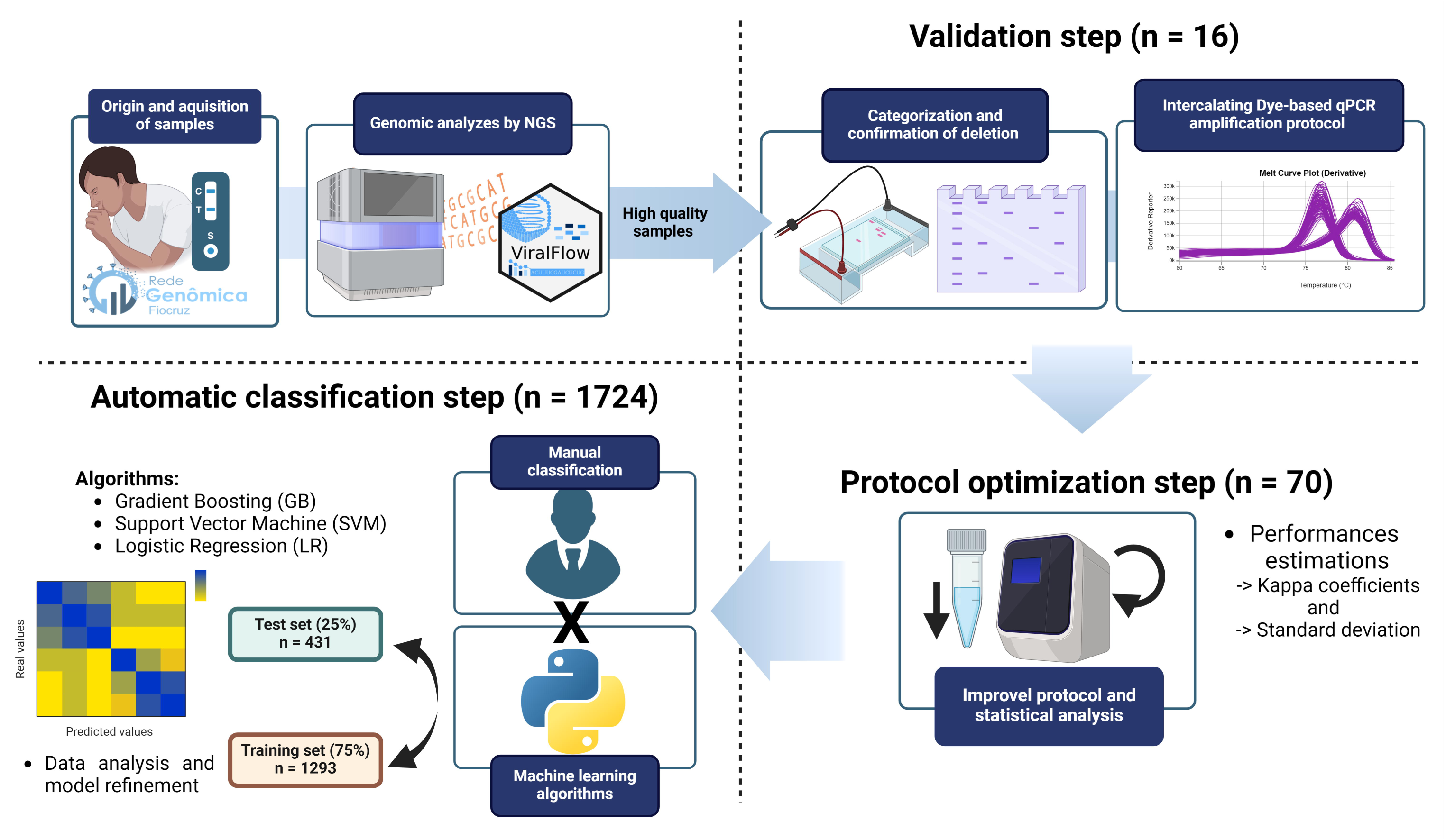
Intercalating dye-based qPCR protocol devised for surveillance of SARS-CoV-2 BE.9 lineages. The schematic illustrates the step-by-step process employed for the precise detection of the targeted deletion within the ORF7a gene (27,508-27,751) by analysis of amplification curves. Synapomorphy was identified by NGS, confirmed through electrophoresis of a subset of high-quality sequencing samples and compared with the amplification results. The protocol was automated using refined machine learning models for an extended set of 1,724 samples, trained through manual classification.

**Figure 2.**
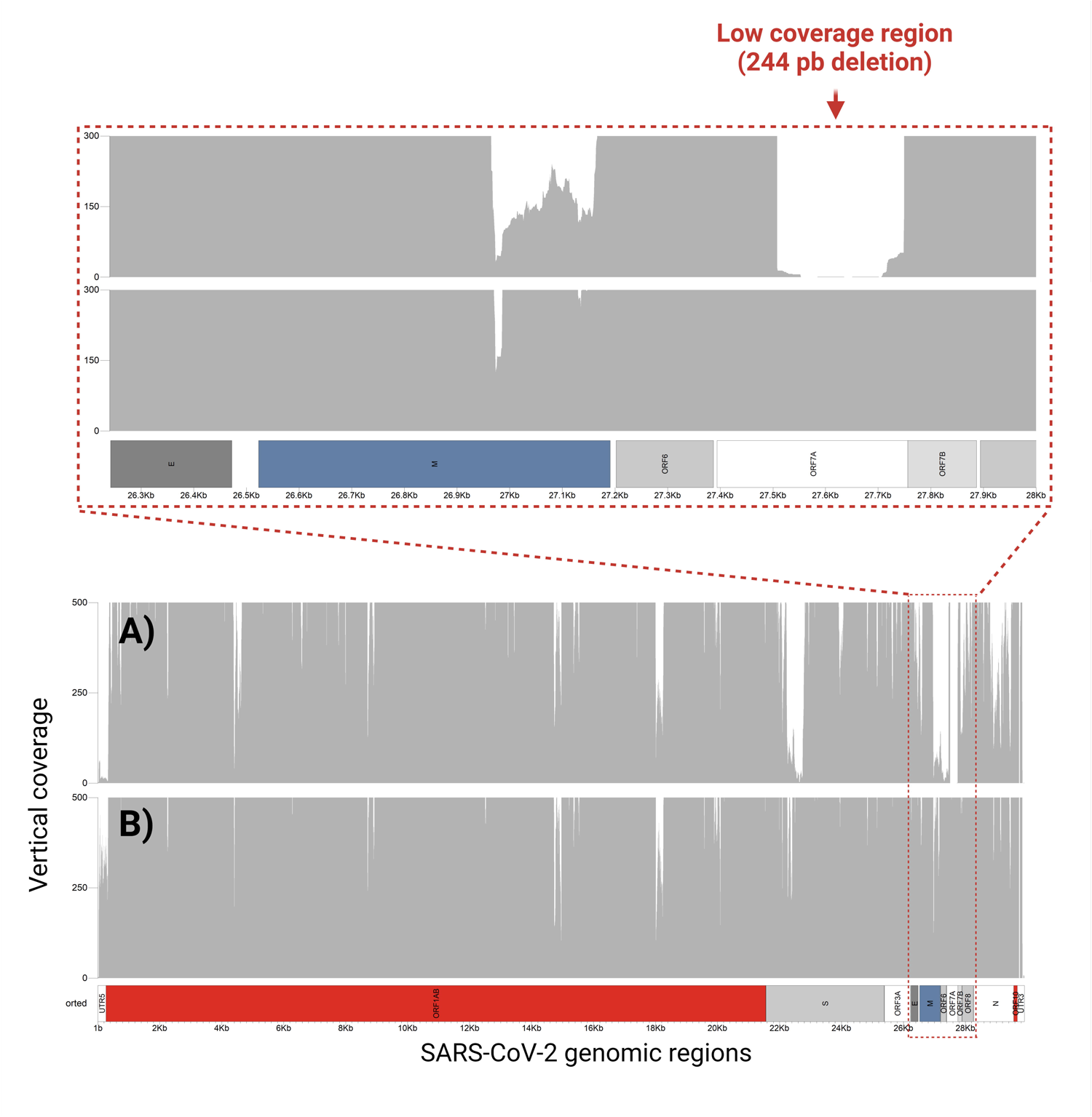
Genomic vertical coverage profile of SARS-CoV-2 highlighting variations. The coverage distribution across the genome shows the difference between BE.9 (A) and non-BE.9 (B) lineages in the ORF7a gene region, showing the low coverage region due to the presence of the 244-base deletion in the BE.9 samples.

The presence of extensive low-depth sequenced regions presents significant challenges to bioinformatics analyses and interpretation, undermining potentially the accurate identification of genuine evolutionary events, such as deletions. The detection of this particular structural mutation (a deletion of 244 bp) within the ORF7a gene was corroborated by routine inspection of amplified targets separated by gel electrophoresis, which endorsed it as a synapomorphic signature of the BE.9 subvariants. The detection of a characteristic band in the range of 170-200 bp (S01 to S08) was found across all BE-9 samples phylogenetically assigned by whole genome sequencing. Amongst the ‘non-BE.9’ samples (S09 to S16), *ORF7a* bands between 400-430 bp were consistently present, thus denoting the absence of deletion **(Figure 3)**. These distinct band patterns offer compelling evidence for the existence of genuine structural alterations between two major SARS-CoV-2 subvariants, of independent origins, and reinforce findings from previous studies concerning the loss of genetic elements during the natural evolution of SARS-CoV-2^3^. Additionally, it also contributed to obtaining evidence regarding the applicability of a PCR amplification protocol that makes use of intercalating dye-based strategies, aiming to increase speediness and robustness of the investigations.

**Figure 3.**
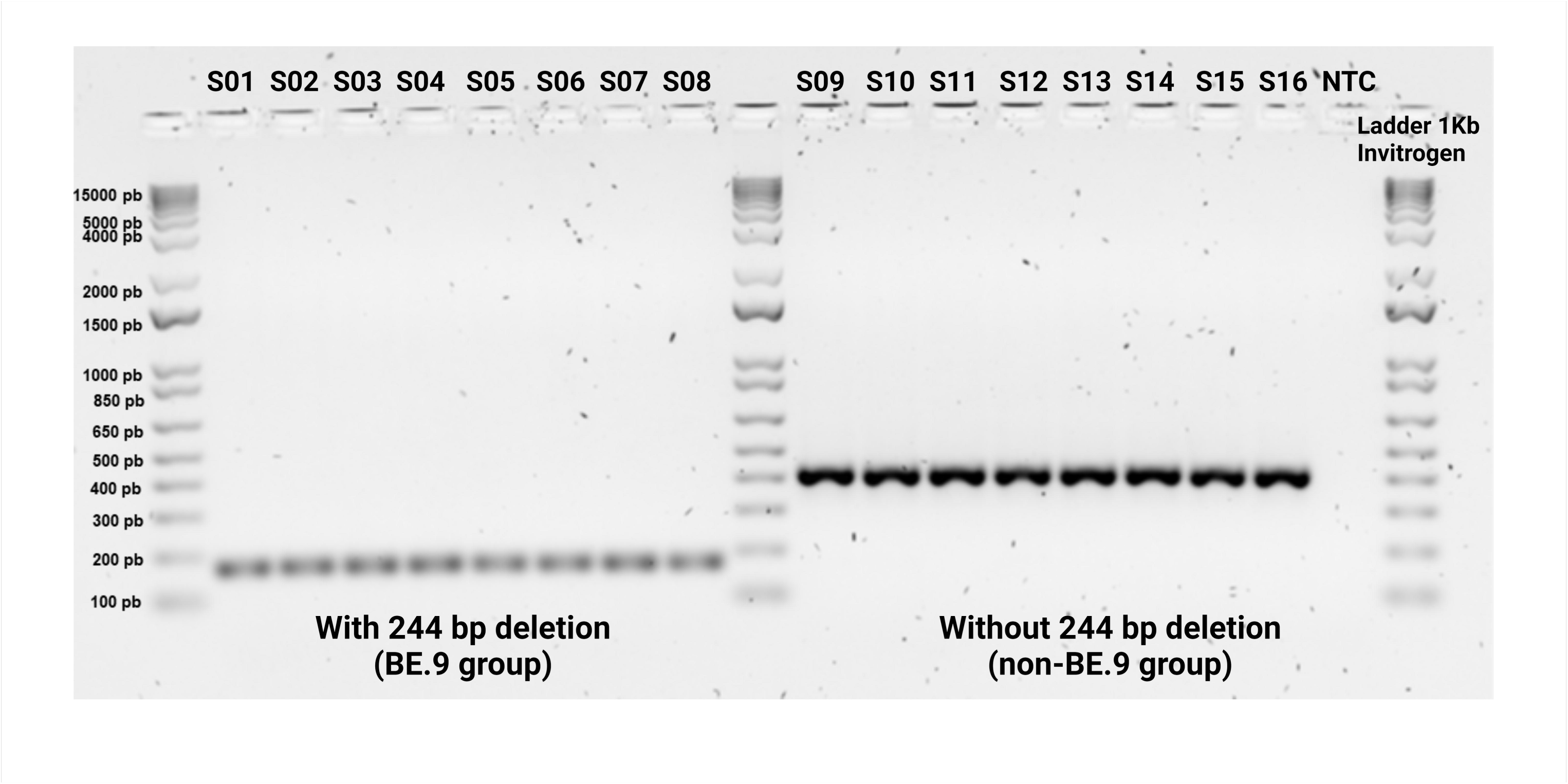
Agarose gel electrophoresis of amplified DNA fragments to validate the ORF7a deletion. The ‘BE.9’ groups (S01 to S08) show the absence of part of the band corresponding to ORF7a, located around 244 base pairs (bp), while the ‘non-BE.9’ groups (S09 to S16) show bands intact sections of ORF7a, between 400 and 430 bp.

### Initial intercalating dye-based qPCR amplification protocol

Building upon these insights, a qPCR protocol enhanced by the integration of the intercalating dye-based assay (BRYT^®^ Green GoTaq mastermix, Promega inc.) aimed into amplifying the region encompassing the identified 244-base pair deletion, thus providing a targeted and high-throughput method for distinguishing between ‘BE.9’ and ‘non-BE.9’ lineages, as well as a reliable, flexible and cost-effective approach ^26,27^.

The results obtained from the first derivative melt curves of the sixteen samples are clarity in **Figure 4**, illustrating the melt curves corresponding to the BE.9 group and the non-BE.9 group. All melt curves for the BE.9 group exhibited amplification at an average melting temperature (TM) of 76.78 ± 0.18°C, accompanied by fluorescence levels ranging between 200k and 300k at its peak. In contrast, the non-BE.9 group displayed an average melting temperature of 80.76 ± 0.24°C, with a wider range of fluorescence intensity, spanning from 200k to 400k. This confirmation, highlighted by the lower Tm for BE.9 and higher Tm for non-BE.9, aligns with prior research suggesting that longer amplicons exhibit higher melting temperatures (TM) compared to shorter ones ^28^. Notably, it was observed during manual analysis and categorization of the samples that the melting curves with fluorescence levels below 100k were challenging to visualize and classify accurately. This challenge was significantly alleviated when the range was filtered to values greater than 100k, enhancing the clarity and precision of group classification.

**Figure 4.**
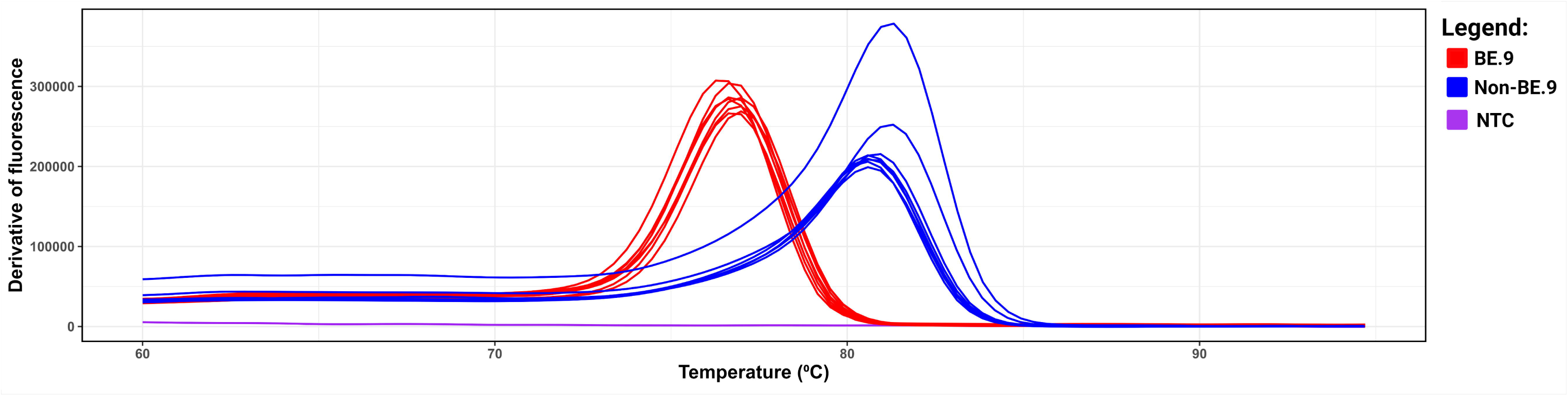
Dissociation curve generated from the ORF7a_244del assay, designed for BE.9 detection via RT-qPCR protocol. This assay targets the 244-base deletion in the ORF7a region of the SARS-CoV-2 genome, a defining characteristic of the BE.9 lineages. Samples attributed to the ‘BE.9’ designation are highlighted in red, consistently exhibiting lower Tm values (76.78 ± 0.18°C), while ‘non-BE.9’ samples, depicted in blue, demonstrate higher Tm values (80.76 ± 0.24°C). Notably, the negative control displayed no amplification.

The first derivative melt graphs of the sixteen samples demonstrated a distinct separation between the BE.9 and non-BE.9 groups. No instances of BE.9 were observed within the melting temperature (TM) range of the non-BE.9 groups, and reciprocally, underscoring the assay’s effectiveness in distinguishing between these virus lineages. The behavior of the negative control (NTC) visualized in **Figure 4**, representing the absence of the virus, exhibited no fluorescence, indicating the absence of primer dimers or unintended products in the assay. This underscores the careful management of primers and ensures the assay’s reliability by minimizing the presence of contaminating artifacts.

### Machine learning algorithms and data analysis

The SVM with a linear kernel emerged as the best-performing model, surpassing Logistic Regression and Gradient Boosting **(Table 2)**.

**Table 2.** Performance Comparison of Machine Learning Algorithms. This table presents the performance metrics of machine learning algorithms tested of different groups: “BE.9”, “non-BE.9”, and “Inconclusive”.

| Algorithm | Precision |  |  | Recall |  |  | F1-score |  |  | Accuracy |
| --- | --- | --- | --- | --- | --- | --- | --- | --- | --- | --- |
|  | BE.9 | Non- BE.9 | Inconclusive | BE.9 | Non-BE.9 | Inconclusive | BE.9 | Non- BE.9 | Inconclusive |  |
| **SVM** | 0.991 | 0.992 | 0.936 | 0.974 | 0.968 | 0.981 | 0.983 | 0.980 | 0.960 | **0.974** |
| **Logistic Regression** | 1.000 | 0.976 | 0.910 | 0.932 | 0.984 | 0.971 | 0.965 | 0.980 | 0.940 | **0.963** |
| **Gradient Boosting** | 0.983 | 0.976 | 0.961 | 0.983 | 0.984 | 0.952 | 0.983 | 0.980 | 0.957 | **0.974** |

Optimizing the SVM parameters resulted in a tie between various hyperparameters configurations **(Supplementary table 3)**. This **Table 3** compares the unoptimized version of the SVM model with the one using the settings {’C’: 100, ‘degree’: 2, ‘kernel’: rbf, gamma: auto} on the evaluate set with the fine-tuned model on the test set. The accuracy demonstrated a marginal improvement, accompanied by enhanced precision, recall, and F1-score metrics for certain subsets within the BE.9, non-BE.9, or inconclusive groups when evaluating the impact of fine-tuning on the test set. This uptick in accuracy indicates the accurate classification of previously mislabeled inconclusive curves as non-BE.9. However, it is imperative to assess these metrics on unseen data. While there was a decrease in metrics, it is probable that these values reflect the true performance on other unseen datasets.

**Table 3.**
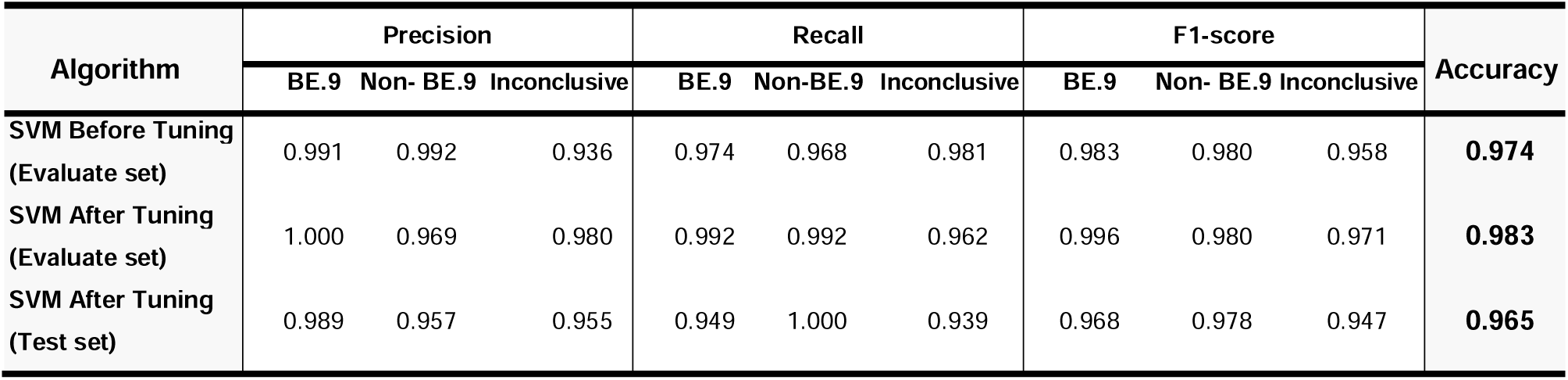
Performance Comparison before and after SVM Parameter Optimization. This table illustrates the performance comparison between the unoptimized version of the Support Vector Machine (SVM) model and the version utilizing specific hyperparameter settings {’C’: 100, ‘degree’: 3, ‘kernel’: ‘linear’}.

| Algorithm | Precision |  |  | Recall |  |  | F1-score |  |  | Accuracy |
| --- | --- | --- | --- | --- | --- | --- | --- | --- | --- | --- |
|  | BE.9 | Non- BE.9 | Inconclusive | BE.9 | Non-BE.9 | Inconclusive | BE.9 | Non- BE.9 | Inconclusive |  |
| SVM Before Tuning<br>(Evaluate set) | 0.991 | 0.992 | 0.936 | 0.974 | 0.968 | 0.981 | 0.983 | 0.980 | 0.958 | **0.974** |
| SVM After Tuning<br>(Evaluate set) | 1.000 | 0.969 | 0.980 | 0.992 | 0.992 | 0.962 | 0.996 | 0.980 | 0.971 | **0.983** |
| SVM After Tuning<br>(Test set) | 0.989 | 0.957 | 0.955 | 0.949 | 1.000 | 0.939 | 0.968 | 0.978 | 0.947 | **0.965** |

The high accuracy signifies substantial reliability when utilizing melting curve points for curve classification, automating the process. Other studies have approached diagnostic classification using derived metrics, whether through Principal Component Analysis (PCA) ^30,31^, metrics related to curve shape (skewness, kurtosis etc) ^20^, or variables associated with the technique itself (amplicon melting temperature) ^32^, achieving accuracies ranging from 72% to 100%. In this work, however, we chose to directly use the curve itself, transforming each point of every curve into a column, or feature, for the model, employing a simple data normalization step. This approach streamlines the model development process, ensuring simplicity without compromising accuracy. It is noteworthy that the clear distinction between curves for BE.9 and non-BE.9 classification enables this approach. By utilizing points from the curve directly, the model gains the flexibility to discern nuances indicating which individual points are more crucial for correct result classification.

The confusion matrix reveals correct classification values for the BE.9 lineage at 94.85% (n = 92), non-BE.9 at 100.00% (n = 134), and inconclusive at 93.86% (n = 107) **(Figure 5)**. Despite the high classification accuracy for SARS-CoV-2 BE.9 and non-BE.9 lineages, there is a noticeable decline in classification quality for inconclusive curves, often reflecting the subjective nature of classification by analysts. It is crucial, therefore, during the establishment of the gold standard used for model training, to clearly define each of the curves.

**Figure 5.**
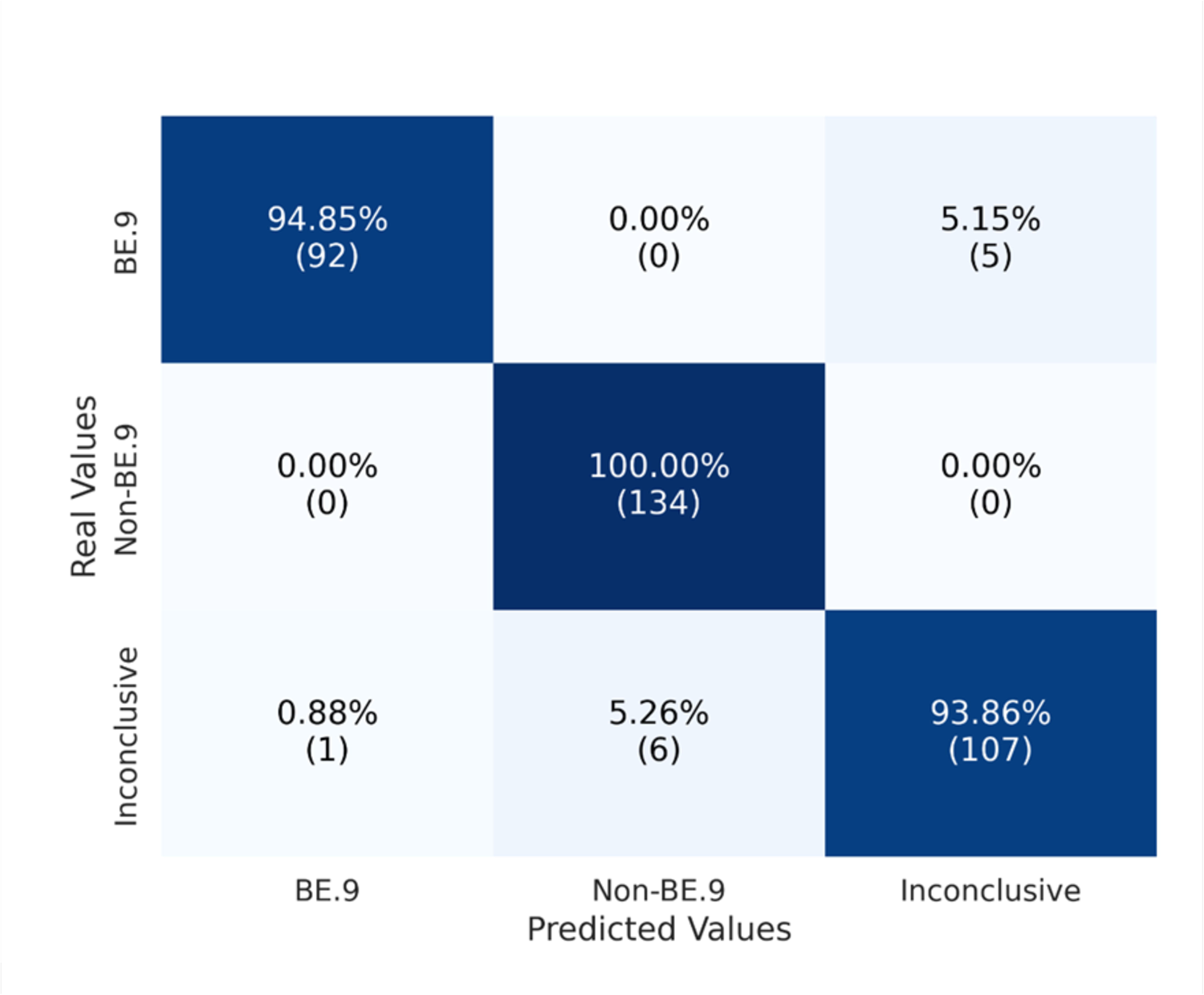
Melting Curve Classification Performance. The performance of melting curve classification shows the high accuracy achieved, indicating substantial reliability in automating the classification process.

The utilization of free platforms such as Google Colaboratory could contribute to the democratization and swift investigation of outbreaks of new variants in regions lacking computational power ^33^. Simple modeling from minimally processed data represents an encouraging opportunity for other groups to optimize protocols, demystifying the use of machine learning algorithms in routine laboratory procedures, allowing for biological applications as already used for other purposes ^34,35^.

## CONCLUSIONS

In conclusion, our study demonstrates the efficacy of the implemented optimized intercalating dye-based qPCR protocol combined with machine learning (ML) analysis as a powerful method for discriminating and classifying independent SARS-CoV-2 sublineages of high homology. This approach offers automated binary inference of the most probable circulating SARS-CoV-2 sublineages (BE.9 or non-BE.9), providing a valuable complement to the more complex NGS-based surveillance methods. The identification of a region of low vertical coverage in BE.9 samples, confirmed through gel electrophoresis as a genuine synapomorphy in the form of a 244 bp deletion, underscores the importance of structural genomic alterations in providing alternatives for monitoring emergence and spread of SARS-CoV-2 variants.

Moreover, the distinct melting temperature (TM) curves between ‘BE.9’ and ‘non-BE.9’ groups, along with a classification sensibility of 94.85% and 100.00%, respectively, using the SVM ML algorithm, highlight the robustness of our methodology. Despite initial challenges with “inconclusive” samples, primarily stemming from characteristics of reused rapid antigen tests, our method maintained a high classification sensibility of 93.86% for identifying such samples. These results underscore the potential of qPCR-based protocols for investigating evolutionary patterns in pathogens, with broad implications for diagnostics, surveillance, and public health interventions.

Moving forward, further research is warranted to validate and refine our method, extending its applicability to other infectious diseases and addressing any existing limitations. This will ensure its continued relevance in the dynamic landscape of infectious disease research and control. Additionally, the integration of machine learning methodology, as demonstrated in this study, enhances the analytical capabilities of generated data, ultimately optimizing lineage diagnosis.

Furthermore, exploring the potential application of non-specific intercalating dye assays for detecting and identifying various pathogens opens avenues for extending this innovative methodology of machine learning to other assays. This broader application not only enhances its utility but also reduces costs and the need for robust equipment, making it more accessible to diverse research settings. Overall, our study contributes to advancing methodologies in infectious disease research and underscores the potential of interdisciplinary approaches in combating emerging pathogens.

## METHODS

### Origin and acquisition of samples

The viral RNA samples were acquired through a collaborative initiative focused on genomic monitoring of SARS-CoV-2, conducted by the Fiocruz Genomic Surveillance Network—an entity under the Brazilian Ministry of Health. These samples were derived from the repurposing of rapid antigen tests conducted as part of routine clinical care, screening processes, and active surveillance for variants in hospitals and health centers in the state of Ceara, Brazil.

### Integrative Genomic Analysis and Categorization

The paired-end sequencing was conducted during the routine process for genomic surveillance of SARS-CoV-2, employing the Artic v4.1 primer set (https://github.com/artic-network/artic-ncov2019/blob/master/primer_schemes/nCoV-2019/V4/SARS-CoV-2.primer.bed) in conjunction with the CovidSeq protocol, used as recommended by the manufacturer, implemented on the Illumina NextSeq 2000 platform for all samples. The raw sequencing data underwent a rigorous analysis utilizing the ViralFlow v1.0.0 workflow (https://viralflow.github.io/), which encompasses quality control, pre-processing, alignment of high-quality reads to the reference genome and genome assembly. Lineage classification was executed utilizing the Pangolin v4.3.1 ^36^ and Nextclade v3.0.1 ^37^ softwares, which facilitated the identification and annotation of genetic variations.

Sixteen high-quality sequencing samples were meticulously chosen for the validation step based on stringent criteria, ensuring horizontal coverage exceeding 90% and vertical coverage surpassing 100x. These samples were drawn from two distinct groups: the ‘BE.9’ group, comprising S01 to S08, and the ‘non-BE.9’ group (other lineages), consisting of S09 to S16, based on lineage classification generated by Pangolin. The BAM file was evaluated using the Geneious prime software and the coverage variation throughout the genome was used to predict the 244 bp deletion present in the ORF7a gene of BE.9 group.

To confirm the presence of the anticipated ORF7a deletion, genomic DNA from each selected sample underwent 2% agarose gel electrophoresis. Electrophoresis was conducted for 4 hours at 90V, facilitating thorough separation and visualization of DNA fragments, including the targeted deletion in ORF7a. The bands were viewed using ThermoFisher iBright equipment, allowing instantaneous image generation.

### Machine learning algorithms and data analysis

A total of 1,724 curves of the optimized protocol were manually analyzed and categorized based on previously established standards as ‘BE.9’, ‘non-BE.9’, or ‘Inconclusive’, and the curve points were served as input for model training. The total curves were separated into a matrix X, containing all points from all curves, and a vector y, containing correct classification values for each curve. The 192nd point of each curve (last column of matrix X) was removed due to 475 samples having a null value at this position. Following, the data was split into training sets (60%, n = 1034), evaluate set (20%, n = 345) and test sets (20%, n = 345). Subsequently, X values were normalized to a range of 0 to 1, crucial for unbiased training of two employed models and the X train was balanced to prevent over representative class bias.

Three machine learning algorithms were employed for data modeling: Gradient Boosting (GB), Support Vector Machine (SVM), and Logistic Regression (LR). Models were run with default parameters, except for SVM, where the ‘kernel’ parameter was changed to ‘linear’ instead of ‘rbf.’ The analysis was conducted using Python 3.10.12 in conjunction with the Scikit-learn 1.4.0 library (for Support Vector Machine and Logistic Regression) and XGBoost 2.0.3 package (for Gradient Boosting), all implemented within the Google Colaboratory environment. The code used for training the machine learning models is available in Supplementary Material 1.

The model, exhibiting the highest accuracy, reflecting overall correctness, underwent a grid search optimization step, exploring different parameters for fine-tuning, with a particular focus on optimizing for the accuracy parameter. The supplementary table 3 compiles the results of the grid search ^38^.

## Data Availability

All data produced in the present work are contained in the manuscript

