## Supplementary Material 1 for "Enhancing SARS-CoV-2 Lineage surveillance through the integration of a simple and direct qPCR-based protocol adaptation with established machine learning algorithms": Supplementary Material 1.ipynb

```
# %% [code]
!python --version

# %% [markdown]
### Code Explanation

#### Library Imports
Libraries such as Pandas, NumPy, and scikit-learn are imported for data manipulation, numerical operations, and machine learning functionalities. Seaborn and Matplotlib are imported for visualization purposes.

#### Data Preprocessing
Assuming there's some data preprocessing steps done before this point such as loading data into a DataFrame and handling missing values.

#### Feature Scaling
MinMaxScaler from scikit-learn is used to scale the features to a range between 0 and 1.

#### Handling Class Imbalance
RandomUnderSampler from imbalanced-learn is used to handle class imbalance by undersampling the majority class.

#### Splitting Data into Train and Test Sets
train_test_split from scikit-learn is used to split the resampled data into training and testing sets.

#### Model Training
A Support Vector Machine (SVM) model is trained using the SVC class from scikit-learn.

#### Model Evaluation Metrics
Precision, recall, F1-score, accuracy, and mean squared error are calculated using appropriate functions from scikit-learn.

#### Hyperparameter Tuning
GridSearchCV from scikit-learn is used for hyperparameter tuning of the SVM model.

#### Visualization
A confusion matrix heatmap is plotted using Seaborn and Matplotlib to visualize the performance of the trained model.


# %% [code]
import pandas as pd
import numpy as np
from collections import Counter
from sklearn.metrics import precision_score
from sklearn.metrics import recall_score
from sklearn.metrics import f1_score
from sklearn.metrics import accuracy_score
import seaborn as sns
import matplotlib.pyplot as plt
from sklearn.svm import SVC
from sklearn.linear_model import LogisticRegression
from sklearn.preprocessing import MinMaxScaler
from sklearn.model_selection import train_test_split
from imblearn.under_sampling import RandomUnderSampler
from sklearn.model_selection import GridSearchCV, StratifiedKFold
from sklearn.metrics import mean_squared_error

# %% [markdown]
#### Reading Data
Two Excel files, "CompiledRuns.xlsx" and "MeltingPoints.xlsx", are read into Pandas DataFrames (`dfResults` and `dfMelting` respectively) using the `pd.read_excel()` function.

#### Filtering Data
1. `dfResults` is filtered to include only rows where the value in the "Run" column is greater than or equal to 6.
2. From the filtered `dfResults`, a new DataFrame `dfResultsFiltered` is created containing only the "ConcatName" and "Result" columns.

#### Data Exploration
1. `dfResultsFiltered.shape`: Returns the shape (number of rows and columns) of the DataFrame `dfResultsFiltered`.
2. `dfResultsFiltered.isnull().sum()`: Returns the sum of missing values in each column of the DataFrame `dfResultsFiltered`.
3. `dfResultsFiltered.value_counts("Result")`: Returns the count of unique values in the "Result" column of the DataFrame `dfResultsFiltered`.

# %% [code]
dfResults = pd.read_excel("CompiledRuns.xlsx",sheet_name="Results")
dfMelting = pd.read_excel("MeltingPoints.xlsx",sheet_name="Points")

# %% [code]
dfResults = dfResults[dfResults['Run'] >=6]

# %% [code]
dfResultsFiltered = dfResults[["ConcatName", "Result"]]

# %% [code]
dfResultsFiltered.shape

# %% [code]
dfResultsFiltered.isnull().sum()

# %% [code]
dfResultsFiltered.value_counts("Result")


# %% [markdown]
### Code Explanation

#### Creating Sequential Observations
A new column named "Observation" is added to the DataFrame `dfMelting`. This column represents the sequential count of observations within each group defined by the "ConcatName" column.

#### Pivot Table
A pivot table is created from the DataFrame `dfMelting`. The index of the pivot table is set to "ConcatName", the columns are set to "Observation", and the values are extracted from the "Derivative" column.

#### Renaming Columns
The columns of the pivot table (`df_pivot`) are renamed using list comprehension, where each column is given a formatted name.


# %% [code]

dfMelting['Observation'] = dfMelting.groupby('ConcatName').cumcount() + 1


df_pivot = dfMelting.pivot(index='ConcatName', columns='Observation', values='Derivative')


df_pivot.columns = [f'{col}' for col in df_pivot.columns];df_pivot

# %% [markdown]
#### Merging DataFrames
Two DataFrames, `dfResultsFiltered` and `df_pivot`, are merged using the `pd.merge()` function. The merge is performed on the "ConcatName" column, with a left join specified using the `how='left'` parameter.


# %% [code]
df_merged = pd.merge(dfResultsFiltered, df_pivot, on='ConcatName', how='left')

# %% [code]
df_merged.head()

# %% [code]
df_merged.shape

# %% [markdown]
#### Checking for Missing Values
The `isnull().sum()` function is used to calculate the sum of missing values in each column of the DataFrame `df_merged`.

#### Filtering Rows with Missing Values
The DataFrame `df_merged` is filtered to include only rows where the value in the column '192' is missing, using the `pd.isna()` function.


# %% [code]
df_merged.isnull().sum()

# %% [code]
df_merged[pd.isna(df_merged['192'])]

# %% [markdown]
#### Dropping Columns
The column labeled '192' is dropped from the DataFrame `df_merged` using the `drop()` function.


# %% [code]
df_merged = df_merged.drop(columns=['192'])

# %% [markdown]
#### Splitting Target and Features
- `y = df_merged['Result']`: Creates a Series `y` containing the target variable 'Result' from the DataFrame `df_merged`.
- `X = df_merged.drop(['ConcatName', 'Result'], axis=1)`: Creates a DataFrame `X` containing the features by dropping the columns 'ConcatName' and 'Result' from `df_merged`.

#### Splitting Data into Train and Test Sets
- `X_train, X_test, y_train, y_test = train_test_split(X, y, random_state=0)`: Splits the features and target variables into training and testing sets. `X_train` and `X_test` contain the feature values for training and testing respectively, while `y_train` and `y_test` contain the corresponding target values. The `random_state=0` parameter ensures reproducibility by fixing the random seed.

#### Feature Scaling
- `scaler = MinMaxScaler()`: Initializes a MinMaxScaler object, which is used to scale features to a specific range (by default, between 0 and 1).
- `X_train_normalized = scaler.fit_transform(X_train)`: Fits the scaler to the training data (`X_train`) and then transforms it to normalized values.
- `X_test_normalized = scaler.transform(X_test)`: Uses the scaler fitted on the training data to transform the test data (`X_test`) to normalized values. It's important to note that the same scaler instance is used for both training and testing sets to ensure consistency in scaling.

# %% [code]
y = df_merged['Result']

X = df_merged.drop(['ConcatName', 'Result'], axis=1)


# %% [code]
# BE.9: 0
# Non-BE.9: 1
# Inconclusive: 2

X_train_eval, X_test, y_train_eval, y_test = train_test_split(
    X, y, random_state=0, test_size=0.20)

X_train, X_eval, y_train, y_eval = train_test_split(
    X_train_eval, y_train_eval, random_state=0, test_size=0.25)


# %% [code]
scaler = MinMaxScaler()

X_train_normalized = scaler.fit_transform(X_train)
X_eval_normalized = scaler.transform(X_eval)
X_test_normalized = scaler.transform(X_test)


# %% [markdown]
#### Resampling with Random Under-sampling
- `under_sampler = RandomUnderSampler(random_state=42)`: Initializes a RandomUnderSampler object with a fixed random state of 42. Random under-sampling is a technique used to balance imbalanced datasets by randomly removing samples from the majority class.
- `X_res_normalized, y_res_normalized = under_sampler.fit_resample(X_train_normalized, y_train)`: Applies random under-sampling to the normalized training data (`X_train_normalized`) and corresponding target variable (`y_train`). The `fit_resample()` method fits the under-sampler to the training data and then applies the resampling technique to balance the classes. The resulting `X_res_normalized` and `y_res_normalized` contain the resampled feature and target data respectively.

# %% [code]

under_sampler = RandomUnderSampler(random_state=42)

X_res_normalized, y_res_normalized = under_sampler.fit_resample(X_train_normalized, y_train)


# %% [code]
print(X_res_normalized.shape)
print(X_eval_normalized.shape)
print(X_test_normalized.shape)

# %% [markdown]
#### Support Vector Machine (SVM)
- `svm_model = SVC(kernel='linear')`: Initializes a Support Vector Machine (SVM) model with a linear kernel. SVM is a supervised learning algorithm used for classification and regression tasks.
- `svm_model.fit(X_res_normalized, y_res_normalized)`: Trains the SVM model on the resampled and normalized training data (`X_res_normalized`) and corresponding target variable (`y_res_normalized`).
- `y_pred_svm = svm_model.predict(X_test_normalized)`: Predicts the target variable for the normalized test data (`X_test_normalized`) using the trained SVM model.

#### Logistic Regression
- `logistic_model = LogisticRegression()`: Initializes a logistic regression model. Logistic regression is a classification algorithm used to model the probability of a outcome.
- `logistic_model.fit(X_res_normalized, y_res_normalized)`: Trains the logistic regression model on the resampled and normalized training data (`X_res_normalized`) and corresponding target variable (`y_res_normalized`).
- `y_pred_reg = logistic_model.predict(X_test_normalized)`: Predicts the target variable for the normalized test data (`X_test_normalized`) using the trained logistic regression model.

#### Gradient Boosting
- `from xgboost import XGBClassifier`: Imports the XGBoost classifier from the XGBoost library. XGBoost is an implementation of gradient boosting algorithms.
- `model = XGBClassifier(n_jobs=-1)`: Initializes an XGBoost classifier with all available CPU cores (`n_jobs=-1`).
- `model.fit(X_res_normalized, y_res_normalized)`: Trains the XGBoost classifier on the resampled and normalized training data (`X_res_normalized`) and corresponding target variable (`y_res_normalized`).
- `y_pred_gb = model.predict(X_test_normalized)`: Predicts the target variable for the normalized test data (`X_test_normalized`) using the trained XGBoost classifier.

# %% [code]
# SVM

svm_model = SVC(kernel='linear')

svm_model.fit(X_res_normalized, y_res_normalized)

y_pred_svm = svm_model.predict(X_eval_normalized)


# %% [code]
# Logistic regression

logistic_model = LogisticRegression()

logistic_model.fit(X_res_normalized, y_res_normalized)

y_pred_reg = logistic_model.predict(X_eval_normalized)


# %% [code]
# GradientBoosting

from xgboost import XGBClassifier

model = XGBClassifier(n_jobs = -1)

model.fit(X_res_normalized, y_res_normalized)

y_pred_gb = model.predict(X_eval_normalized)


# %% [markdown]
#### Support Vector Machine (SVM) Metrics
- `print('Precision score %s' % precision_score(y_test, y_pred_svm, average=None))`: Computes and prints the precision score for each class separately using the `precision_score` function.
- `print('Recall score %s' % recall_score(y_test, y_pred_svm, average=None))`: Computes and prints the recall score for each class separately using the `recall_score` function.
- `print('F1-score score %s' % f1_score(y_test, y_pred_svm, average=None))`: Computes and prints the F1-score for each class separately using the `f1_score` function.
- `print('Accuracy score %s' % accuracy_score(y_test, y_pred_svm))`: Computes and prints the accuracy score using the `accuracy_score` function.

#### Logistic Regression Metrics
- Similar to SVM, these lines of code compute and print precision, recall, F1-score, and accuracy metrics for the logistic regression model (`y_pred_reg`).

#### Gradient Boosting Metrics
- Similar to SVM and logistic regression, these lines of code compute and print precision, recall, F1-score, and accuracy metrics for the gradient boosting model (`y_pred_gb`).

# %% [code]
# SVM

print('Precision score %s' % precision_score(y_eval, y_pred_svm, average=None))
print('Recall score %s' % recall_score(y_eval, y_pred_svm, average=None))
print('F1-score score %s' % f1_score(y_eval, y_pred_svm, average=None))
print('Accuracy score %s' % accuracy_score(y_eval, y_pred_svm))

# %% [code]
# Logistic Regression

print('Precision score %s' % precision_score(y_eval, y_pred_reg, average=None))
print('Recall score %s' % recall_score(y_eval, y_pred_reg, average=None))
print('F1-score score %s' % f1_score(y_eval, y_pred_reg, average=None))
print('Accuracy score %s' % accuracy_score(y_eval, y_pred_reg))

# %% [code]
# GradientBoosting

print('Precision score %s' % precision_score(y_eval, y_pred_gb, average=None))
print('Recall score %s' % recall_score(y_eval, y_pred_gb, average=None))
print('F1-score score %s' % f1_score(y_eval, y_pred_gb, average=None))
print('Accuracy score %s' % accuracy_score(y_eval, y_pred_gb))

# %% [markdown]
#### Results Dictionary
A dictionary named `results` is created to store the evaluation metrics for different algorithms. Each key in the dictionary corresponds to a different metric, and the values are lists containing the metric values for each algorithm.

#### Creating DataFrame
The dictionary `results` is used to create a Pandas DataFrame named `df_results`. Each key in the dictionary becomes a column in the DataFrame.

#### Saving to File
The DataFrame `df_results` is saved to a tab-separated values (TSV) file named "AlgorithmComparison.tsv" using the `to_csv()` function. The parameter `sep='\t'` specifies that tabs should be used as separators between values, and `index=False` ensures that the DataFrame index is not included in the output file.

# %% [code]

results = {
    'Algorithm': ['SVM', 'Logistic Regression', 'GradientBoosting'],
    'Precision': [
        precision_score(y_eval, y_pred_svm, average='micro'),
        precision_score(y_eval, y_pred_reg, average='micro'),
        precision_score(y_eval, y_pred_gb, average='micro')
    ],
    'Recall': [
        recall_score(y_eval, y_pred_svm, average='micro'),
        recall_score(y_eval, y_pred_reg, average='micro'),
        recall_score(y_eval, y_pred_gb, average='micro')
    ],
    'F1-score': [
        f1_score(y_eval, y_pred_svm, average='micro'),
        f1_score(y_eval, y_pred_reg, average='micro'),
        f1_score(y_eval, y_pred_gb, average='micro')
    ],
    'Accuracy': [
        accuracy_score(y_eval, y_pred_svm),
        accuracy_score(y_eval, y_pred_reg),
        accuracy_score(y_eval, y_pred_gb)
    ]
}

df_results = pd.DataFrame(results)


df_results.to_csv('AlgorithmComparison.tsv',sep='\t',index=False)

# %% [markdown]
#### Function Definition
- The function `plotMatrix` is defined to visualize the confusion matrix for classification models.
- It takes three arguments: `y_pred` (predicted labels), `labels` (list of class labels, defaulting to ["BE.9", "Non-BE.9", "Inconclusive"]), and `figName` (name of the figure file to be saved, defaulting to "ConfusionMatrix").

#### Confusion Matrix Computation
- Inside the function, the confusion matrix is computed using the predicted labels (`y_pred`) and the actual labels (`y_test`).
- The confusion matrix is then converted to percentages by dividing each element by the sum of its corresponding row.

#### Plot Configuration
- The function configures the heatmap plot using Seaborn and Matplotlib libraries.
- For each cell in the heatmap, text is added to display the percentage and count of samples, with text color set based on the percentage value.

#### Saving and Displaying Plot
- The plot is saved as a PNG image with the filename specified by `figName`.
- The plot is displayed using Matplotlib's `plt.show()` function.


# %% [code]
import numpy as np
import seaborn as sns
import matplotlib.pyplot as plt
from sklearn.metrics import confusion_matrix

def plotMatrix(y_test, y_pred, labels = ["BE.9", "Non-BE.9", "Inconclusive"], figName = "ConfusionMatrix"):

  conf_matrix = confusion_matrix(y_test, y_pred)
  conf_matrix_percent = conf_matrix / np.sum(conf_matrix, axis=1, keepdims=True)

  # Configuração do gráfico
  class_names = labels
  sns.set(font_scale=1.2)
  plt.figure(figsize=(8, 6))
  sns.heatmap(conf_matrix_percent, annot=False, fmt="d", cmap="Blues", xticklabels=class_names, yticklabels=class_names, cbar=False)
  for i in range(len(class_names)):
      for j in range(len(class_names)):
          if conf_matrix_percent[i, j] > 0.7:
            color = "white"
          else:
            color="black"
          plt.text(j + 0.5, i + 0.5, f"{conf_matrix_percent[i, j]:.2%}\n({conf_matrix[i, j]})", ha="center", va="center", color=color)

  plt.xlabel('Predicted Values')
  plt.ylabel('Real Values')
  plt.savefig(f'{figName}.png', dpi=300)
  plt.show()

# %% [code]
plotMatrix(y_eval, y_pred_svm, figName = "svmMatrix")

# %% [markdown]
#### Imports
- The necessary libraries such as Pandas, SVC (Support Vector Classifier) from scikit-learn, GridSearchCV for hyperparameter tuning, and various metrics from scikit-learn are imported.

#### Creating Results DataFrame
- A DataFrame named `results_df` is created to store the results of different SVM models with various hyperparameters. It includes columns for parameters, mean squared error (MSE), precision, recall, F1-score, and accuracy.

#### Default SVM Model
- A default SVM model (`svm_model_standard`) is trained using the default parameters and evaluated on the test set to obtain baseline performance metrics such as MSE, precision, recall, F1-score, and accuracy.

#### Parameter Grid
- A parameter grid (`param_grid`) is defined, specifying various values for the hyperparameters 'C' (regularization parameter), 'kernel' (type of kernel function), 'gamma' (kernel coefficient), and 'degree' (degree of the polynomial kernel).

#### Grid Search
- GridSearchCV is used to perform hyperparameter tuning for the SVM model. It searches through different combinations of hyperparameters specified in `param_grid` to find the combination that optimizes the accuracy metric.

#### Model Evaluation
- For each combination of hyperparameters tested during the grid search, a new SVM model is trained using those parameters, and its performance is evaluated on the test set. Metrics such as MSE, precision, recall, F1-score, and accuracy are computed for each model.

#### Saving Results
- The results, including hyperparameters and evaluation metrics, are stored in the DataFrame `results_df`.
- The DataFrame is saved to an Excel file named "results_svm.xlsx" using the `to_excel()` function for further analysis.

# %% [code]

import pandas as pd
from sklearn.svm import SVC
from sklearn.model_selection import GridSearchCV
from sklearn.metrics import mean_squared_error, precision_score, recall_score, f1_score, accuracy_score
from sklearn.model_selection import StratifiedKFold

# Creating a DataFrame to store the results
results_df = pd.DataFrame(columns=['Parameters', 'Precision', 'Recall', 'F1', 'Accuracy'])

# Default SVM Configuration
svm_model_standard = SVC()
svm_model_standard.fit(X_res_normalized, y_res_normalized)
y_pred_standard = svm_model_standard.predict(X_eval_normalized)

# Evaluating the default model on the test set
precision_standard = precision_score(y_eval, y_pred_standard, average='micro')
recall_standard = recall_score(y_eval, y_pred_standard, average='micro')
f1_standard = f1_score(y_eval, y_pred_standard, average='micro')
accuracy_standard = accuracy_score(y_eval, y_pred_standard)

# Adding default results to the DataFrame
results_df = results_df.append({
    'Parameters': 'Default',
    'Precision': precision_standard,
    'Recall': recall_standard,
    'F1': f1_standard,
    'Accuracy': accuracy_standard
}, ignore_index=True)

# Defining the parameters you want to test
param_grid = {
    'C': [0.001, 0.01, 0.1, 1, 10, 100, 1000],
    'kernel': ['linear', 'poly', 'rbf'],
    'gamma': [0.01, 0.1, 1, 'auto'],
    'degree': [2, 3, 4],
}

# Creating a GridSearchCV object
grid_search = GridSearchCV(SVC(), param_grid, scoring='accuracy', cv=StratifiedKFold(n_splits=5), verbose=1, n_jobs=-1)

# Running the grid search
grid_search.fit(X_res_normalized, y_res_normalized)

# Iterating over all tested parameter combinations
for i, params in enumerate(grid_search.cv_results_['params']):
    # Creating an instance of the SVC model with the best parameters
    best_model = SVC(**params)

    # Training the model with the best parameters
    best_model.fit(X_res_normalized, y_res_normalized)

    # Evaluating the best model on the test set
    y_pred = best_model.predict(X_eval_normalized)
    precision = precision_score(y_eval, y_pred, average='micro')
    recall = recall_score(y_eval, y_pred, average='micro')
    f1 = f1_score(y_eval, y_pred, average='micro')
    accuracy = accuracy_score(y_eval, y_pred)

    # Adding grid search results to the DataFrame
    results_df = results_df.append({
        'Parameters': params,
        'Precision': precision,
        'Recall': recall,
        'F1': f1,
        'Accuracy': accuracy
    }, ignore_index=True)

# Saving the results to an Excel file
results_df.to_excel('results_svm.xlsx', index=False)


# %% [markdown]
#### Training the Model
- The code begins by training the SVM model with the best hyperparameters.
- An SVM model for classification is created with the following hyperparameters:
  - Kernel: Radial Basis Function (RBF), specified by `kernel='rbf'`.
  - Gamma: Auto, specified by `gamma="auto"`. Gamma defines how far the influence of a single training example reaches, with a higher value indicating a tighter fit.
  - Degree: 2, specified by `degree=2`. Degree is the degree of the polynomial kernel function (only relevant if `kernel='poly'`).
  - Regularization Parameter (C): 100, specified by `C=100`. C controls the trade-off between a smooth decision boundary and classifying the training points correctly.
- The created SVM model is stored in the variable `svm_modelTunned`.

#### Model Training
- The created SVM model (`svm_modelTunned`) is trained on the normalized and resampled training data (`X_res_normalized`, `y_res_normalized`) using the `fit()` method.

#### Making Predictions
- Once the model is trained, predictions are made on the normalized test set (`X_test_normalized`) using the `predict()` method.
- The predicted labels are stored in the variable `y_pred_svmTunned`.

# %% [code]
# SVM

# Training the model with the best hyperparameters
# Creating an SVM model for classification
svm_modelTunned = SVC(kernel='rbf', gamma="auto", degree=2, C=100)
# Training the model
svm_modelTunned.fit(X_res_normalized, y_res_normalized)

# Making predictions on the eval set
y_pred_eval_svmTunned = svm_modelTunned.predict(X_eval_normalized)

# Making predictions on the test set
y_pred_svmTunned = svm_modelTunned.predict(X_test_normalized)


# %% [code]
# SVM

print('Precision score %s' % precision_score(y_eval, y_pred_eval_svmTunned, average=None))
print('Recall score %s' % recall_score(y_eval, y_pred_eval_svmTunned, average=None))
print('F1-score score %s' % f1_score(y_eval, y_pred_eval_svmTunned, average=None))
print('Accuracy score %s' % accuracy_score(y_eval, y_pred_eval_svmTunned))

# %% [code]
# SVM

print('Precision score %s' % precision_score(y_test, y_pred_svmTunned, average=None))
print('Recall score %s' % recall_score(y_test, y_pred_svmTunned, average=None))
print('F1-score score %s' % f1_score(y_test, y_pred_svmTunned, average=None))
print('Accuracy score %s' % accuracy_score(y_test, y_pred_svmTunned))

# %% [code]
plotMatrix(y_test, y_pred_svmTunned, figName="fineTunnedSVM")

# %% [markdown]
#### Saving the Model
- The `joblib.dump()` function is used to save the trained model.
- The model to be saved is specified as the first argument (`model`).
- The second argument is the file path where the model will be saved (`'/content/drive/MyDrive/Machine Learning/TR/model.joblib'`).
- This file path should include the desired location and filename for the saved model.

#### Note
- It's essential to ensure that the directory specified for saving the model exists and that the file path is accessible.

# %% [code]
import joblib

# Saving the model
joblib.dump(model, '/content/model.joblib')
```
